# Evaluating the effectiveness of the D1 Now intervention to improve outcomes among young adults with type 1 diabetes: Protocol for a cluster randomised controlled trial

**DOI:** 10.1101/2025.11.11.25339994

**Authors:** Aswathi Surendran, Molly Byrne, Eimear Morrissey, Alberto Alvarez-Iglesias, Paddy Gillespie, Anna Hobbins, Duygu Sezgin, Seamus Sreenan, Declan Devane, Cameron Keighron, Michelle Lowry, Marleen Kunneman, Anne M Doherty, Site Collaborative Group, Sean F Dinneen

## Abstract

**Background:** Young adults living with type 1 diabetes (T1D) often experience sub-optimal outcomes due to competing life demands, disruptions in care, reduced clinic attendance, and difficulties in self-management. To improve outcomes among this population, we developed and piloted the D1 Now intervention using a user-centred and theory-informed approach. This protocol describes a cluster randomised controlled trial (RCT) to test the effectiveness and cost-effectiveness of the D1 Now intervention.

**Methods:** A cluster RCT seeking to recruit 348 young adults (aged 18–25) with T1D from 12 hospital diabetes centres in Ireland. Centres will be randomised to receive either standard care or the D1 Now intervention, which includes two components: an agenda-setting tool and a support worker. The primary outcome is the change in HbA1c from baseline to 12-month follow-up. Secondary outcomes include patient-reported psychosocial outcomes, clinical outcomes, self-management outcomes and healthcare utilisation. We will collect data through blood samples, online patient surveys, and patient records at baseline and 12 months. Additionally, we will conduct a cost-effectiveness evaluation and a mixed-methods process evaluation.

**Discussion:** We anticipate that the D1 Now intervention will be both effective and cost-effective in improving clinical and psychosocial outcomes for young adults compared to standard care. The findings from the process evaluation will shed light on how the intervention works (or not) and how implementation into health services (if warranted) can be optimised. If effective, D1 Now will offer a sustainable model of care to support engagement and self-management for young adults living with T1D.

**Trial registration:** ISRCTN Identifier: <u>ISRCTN28944606</u>. Date applied 01 May 2025

## Introduction

Young adulthood is a critical developmental period characterised by significant transitions in lifestyle, education, and healthcare responsibilities. For individuals with type 1 diabetes (T1D), this phase - often referred to as "emerging adulthood" -is associated with increased challenges with self-management and poorer clinical outcomes, due to competing life demands, reduced parental oversight, and physiological changes.^1–4^

During this time, young adults frequently experience high blood glucose levels, heightened risk for diabetic ketoacidosis, and early onset of diabetes-related complications.^5–8^ These challenges are compounded by physiological factors, such as insulin resistance, and lifestyle factors, as well as risk-taking behaviours and peer influences.^2,9,10^ Young adults with T1D are at a heightened risk for depression and diabetes-related distress in comparison to other age groups with T1D and their peers without T1D; these psychological challenges are also associated with higher blood glucose levels.^3^ Mortality rates in this age group are notably higher than in their peers without diabetes.^7,11^

The transition from paediatric to adult healthcare services exacerbates these challenges, as young adults must adapt to new healthcare settings and increased personal responsibility. Traditional paediatric care focuses on family-based support, while adult care shifts towards an individualised approach, often resulting in perceived decreases in care quality and continuity.^5,12^ Research highlights the importance of supportive and continuous relationships between young people and healthcare providers in facilitating smoother transitions and improving diabetes management outcomes.^1,4,5,8,13^

Little is known about what interventions are effective in improving outcomes among young adults with T1D, and the quality of existing studies is often poor.^6^ In response to this dearth of research, the D1 Now intervention was developed to improve outcomes among young adults with T1D by supporting self-management and clinic engagement.^14^ The intervention was developed systematically, adopting a user-centred approach and guided by psychological theory, informed by a systematic review, qualitative research, and expert consensus.^14–18^

### D1 Now Pilot study

The feasibility and acceptability of the D1 Now intervention were evaluated in a pilot cluster-randomised controlled trial (cRCT) involving four young adult diabetes clinics in Ireland.^19,20^ Initially, the D1 Now intervention consisted of three components: an agenda-setting tool, an interactive messaging system and a support worker. The agenda-setting tool was adapted from the Type 1 Consultation (T1C) tool to suit the needs and context of young adults in Ireland.^15,21^ The pilot study demonstrated that the intervention was both feasible and acceptable to young adults with T1D, healthcare providers, and clinic systems – in particular, the agenda-setting tool and the support worker.^15,20,22^

The pilot study confirmed that, with a modification to the D1 Now intervention (removal of the interactive messaging system), it was appropriate to progress to a definitive trial to assess the intervention’s effectiveness, cost-effectiveness, and implementation potential at scale.^20,22^

### D1 Now intervention components

The D1 Now intervention includes two main components: an agenda-setting tool and a support worker.

1. Agenda-setting tool: This is a shared decision-making tool used by young adults and healthcare professionals to identify topics to discuss during clinic visits. The tool has two parts: the first is completed by the young adult in the waiting room. It covers focused discussion topics relevant to young adults, such as personal concerns, hospital admissions, blood glucose levels, confidence in carbohydrate counting, and emotional well-being related to diabetes management. The second is completed during the consultation with the clinician(s). It consists of a goal-setting exercise where young adults identify, discuss, and agree on realistic and achievable self-management goals with healthcare providers. The tool also includes a screening questionnaire for diabetes distress (See supplementary files).^15,20^
2. Support Worker: This is a person in addition to the existing diabetes team who acts as an advocate for young adults and is available before, during and after clinic appointments to provide ongoing support, facilitate communication, and assist with self-management.^23^

A logic model (including the intervention components, their constituent behaviour change techniques, with the proposed mechanisms of impact and outcomes) of how the D1 Now intervention is expected to work is shown in Figure 1.

**Figure 1:**
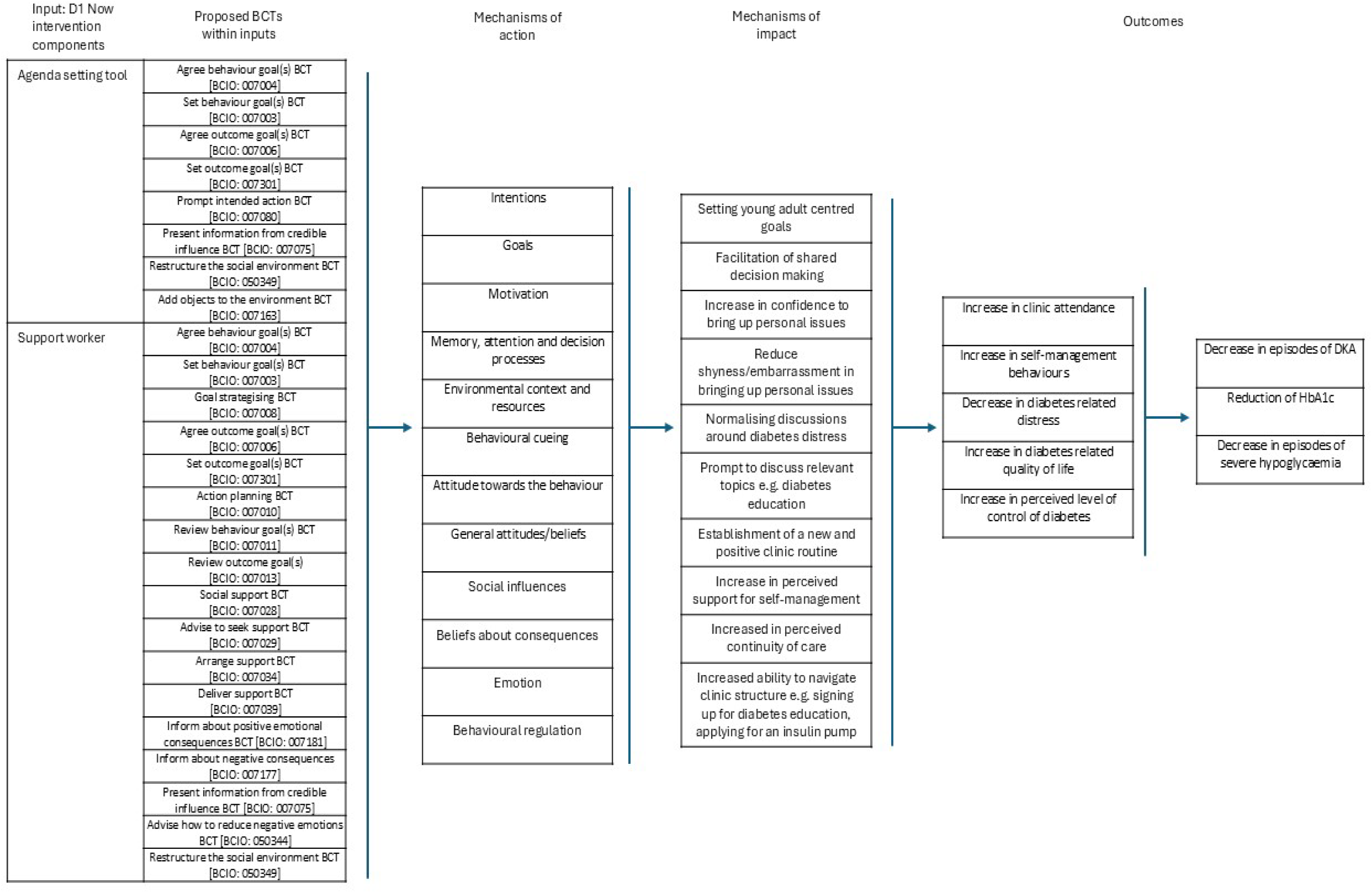
Logic model for how D1 Now is expected to work

### D1 Now Young Adult Panel

The D1 Now Young Adult Panel (YAP), a dedicated Public and Patient Involvement (PPI) group, has been embedded within the research team since the program’s inception. Consisting of up to 10 young adults living with T1D, YAP members have contributed to discussions and decisions around study design, choice of outcomes, design of study materials, and helped design a Study Within a Trial (SWAT) for the Pilot and for the Definitive RCT.^24,25^ They have presented the D1 Now research at conferences, contributed to national policy papers, and supported the recruitment of researchers onto the research team. They continue to be actively involved in trial-related activities, and while new members are regularly recruited to ensure continuity and relevance, several original panel members, now beyond the 18–25 participant age range, remain involved in mentoring roles for incoming members.

### Objectives of the D1 Now Cluster RCT

The primary objective of this study is to evaluate the effectiveness of the D1 Now intervention in reducing HbA1c levels among young adults with T1D compared to standard care.

### Secondary Objectives

1. Evaluate the effectiveness of the D1 Now intervention on secondary clinical outcomes (including young adult clinic attendance, episodes of diabetic ketoacidosis (DKA) and episodes of severe hypoglycaemia) and patient-reported psychosocial outcomes (including diabetes-related distress, self-management behaviours, perceived control over diabetes, diabetes-related quality of life, and diabetes-related stigma).
2. Examine the implementation and delivery of the D1 Now intervention, as well as the overall trial processes.
3. Explore participants’ perceptions and experiences with the D1 Now intervention and standard care.
4. Evaluate the cost-effectiveness of the D1 Now intervention.,

This study will be conducted, analysed, and reported according to the Consolidated Standards of Reporting Trials (CONSORT) 2025 statement extension for cluster RCTs.^27^ Ethical approval will be sought from the relevant Research ethics committees.

## Methods

### Research Design

This study is a multicentre, two-arm, parallel group, cluster RCT across 12 young adult diabetes clinics in Ireland. Clinics will be randomised 1:1 to either D1 Now intervention or standard care.

The flow chart of the study is shown in Figure 2. This protocol is reported in accordance with the Standard Protocol Items: Recommendations for Interventional Trials (SPIRIT) reporting guidelines (see Supplementary file for the SPIRIT Checklist).^28^

**Figure 2:**
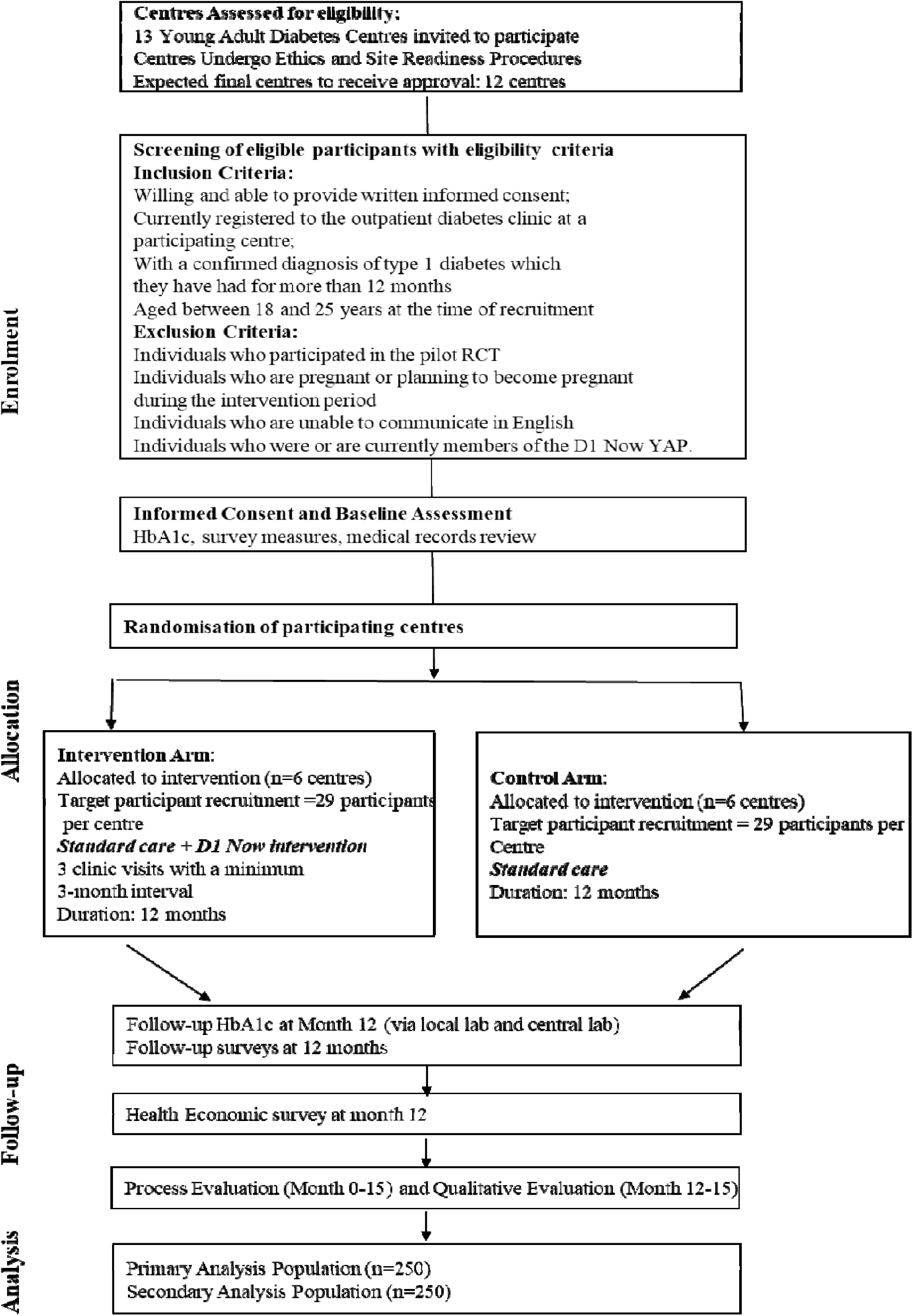
CONSORT statement flow diagram

### Research participants

We aim to recruit 348 young adults (aged 18–25) with T1D from 12 participating clinics (29 participants/clinic). A participant flow diagram is shown in Figure 2.

## Inclusion Criteria for Centres and Participants

### Centres

Included centres (1) operate a dedicated young adult diabetes clinic; (2) have a registered population of at least 60 young adults in the 18-25 age group. This threshold is based on pilot recruitment rates and is considered the minimum required to achieve centre-level enrolment targets, and (3) have a minimum core staff team to include (full or part-time): a Consultant Endocrinologist, a Diabetes Specialist Nurse (DSN) and a Diabetes Specialist Dietitian.

### Participants

Included young adult participants (1) are willing and able to provide written informed consent; (2) are currently registered to the outpatient diabetes clinic at a participating centre; (3) have a confirmed diagnosis of type 1 diabetes for at least the previous 12 months; and (4) are between 18 and 25 years of age at the time of recruitment.

### Exclusion Criteria

#### Participants

Young adult participants who meet any of the following exclusion criteria will not be eligible to take part in the trial: Individuals who (1) participated in the pilot RCT; (2) were or are currently members of the D1 Now Young Adult Panel; (3) are pregnant or planning to become pregnant during the intervention period; and (4) are unable to communicate in English.

### Recruitment

We have identified 13 centres in the Republic of Ireland that meet the eligibility criteria.

We will identify potentially eligible participants through a systematic review of patient registries and medical records at participating sites. Identified individuals will receive the Participant Information Leaflet (PIL) by post or email, along with a link to the online consent form. We will encourage local sites to recruit those young adults who are not regular attenders at outpatient clinics and who may receive much of their diabetes care through unscheduled visits in the Emergency Department and in the hospital. All participants will be given at least two weeks to consider their participation. Once consent is obtained, final eligibility will be confirmed by a study coordinator at each site, and participants will be enrolled into the study.

### Randomisation and Allocation

Diabetes clinics will serve as the unit of randomisation to minimise the risk of contamination between intervention and control conditions. An independent statistician will randomise sites to either the intervention or control arm, using a computer-generated allocation sequence stratified by clinic size. To prevent selection bias, we will randomise sites following participant identification and baseline data collection.

We will employ a simple randomisation in a 1:1 ratio, using randomly permuted blocks of sizes 2 and 4. We will stratify randomisation by the centre’s hospital model (categorised by the Health Service Executive as models 2 (local), 3 (major) or 4 (regional)) to ensure a greater balance between treatment groups. We will generate a single sequence of random assignments for each stratum before treatment allocation starts, using a fixed starting seed for reproducibility.

Allocation concealment will be ensured by having the independent statistician communicate randomisation results directly to sites only after baseline data collection is complete for all enrolled participants at that site.

The nature of the intervention means it is not feasible to blind study coordinators, clinics, participants or data collectors to treatment allocation. However, the primary outcome (HbA1c) will be analysed by laboratory staff blinded to allocation. Statistical analysts will remain blinded to group allocation until the primary analysis is complete, with groups coded as ‘A’ and ‘B’ during analysis.

### Intervention

#### Intervention arm

We will implement the D1 Now intervention as an add-on to standard care in intervention arm centres. The intervention does not require additional clinic visits but aims for at least three clinic appointments over the 12-month follow-up period. Each of these visits will incorporate the use of the agenda-setting tool, plus support from a support worker.

Support is tailored to participant preference and maintained between visits via in-person or virtual communication.

Further detail of the three clinic appointments is provided below.

#### First Clinic Visit (Month 3–4)

On the day of the clinic visit, the young adult will complete part 1 of the agenda-setting tool, with the help of the support worker, before meeting members of the clinic team. The consultation will then proceed using the agenda-setting tool to guide discussions. If the participant’s score is elevated on the Diabetes Distress Scale-2 item (DDS-2)^29^, the clinician can ask them to complete the more comprehensive 28-item version of this scale (T1-DDS-28).^30^ In cases of elevated diabetes distress, the clinician will direct the young adult to locally available resources, based on clinical guidelines for managing diabetes distress. After the consultation, the young adult will meet with the support worker to discuss the appointment, receive a copy of their agenda-setting tool, and agree on any necessary follow-up before the next clinic appointment.

#### Second Clinic Visit (Month 7–8)

The same procedures are repeated. The agenda-setting tool is updated, distress screening is conducted, and the support worker helps with reflection and follow-up.

#### Third Clinic Visit (Month 12)

The procedures will be similar to the previous appointments, with the addition of a follow-up HbA1c blood sample collected on site. Participants will complete the remaining questionnaires online within one month. After the final appointment, the support worker will discharge the young adult from their care and transfer responsibility back to the clinical team.

#### Control Arm

Participants in the control arm will receive standard care as defined by the Irish national clinical guidelines.^1^ Control arm participants will attend regular clinic appointments as scheduled by their care team over the 12-month trial period. More details regarding the standard care will be published in the process evaluation manuscript.

All staff in both arms will receive standardised training on trial procedures, including participant recruitment, data collection, REDCap use, and, where applicable, procedures for coordinating with Support Workers and intervention delivery, to ensure consistency and protocol adherence.

A comprehensive study design and schedule of visits is available in the supplementary file.

### Data collection and outcomes

We will collect research data at two key time points: baseline and 12 months after baseline. The study coordinator, trained in standard operating procedures, will oversee all data collection activities to ensure consistency across participating centres. Following informed consent, participants in both study arms will complete baseline questionnaires online and provide a blood sample during a scheduled clinic visit for HbA1c analysis. These samples will be processed at a central laboratory to ensure consistency across sites.

At the 12-month follow-up, which aligns with the participant’s third clinic visit, both study arms will again complete follow-up questionnaires and provide a second blood sample for HbA1c analysis. The questionnaires will be completed online within one month of the clinic visit. Local site staff will draw the blood samples, and the study coordinator will arrange transport to the central laboratory. Throughout the trial, the study coordinator will also collect implementation and fidelity-related data to support the process evaluation.

Outcomes and their corresponding measurement tools are summarised in Table 1. These were selected based on a core outcome set developed for studies involving young adults with type 1 diabetes.^32^ Full outcome descriptions are available in the supplementary materials.

**Table 1:**
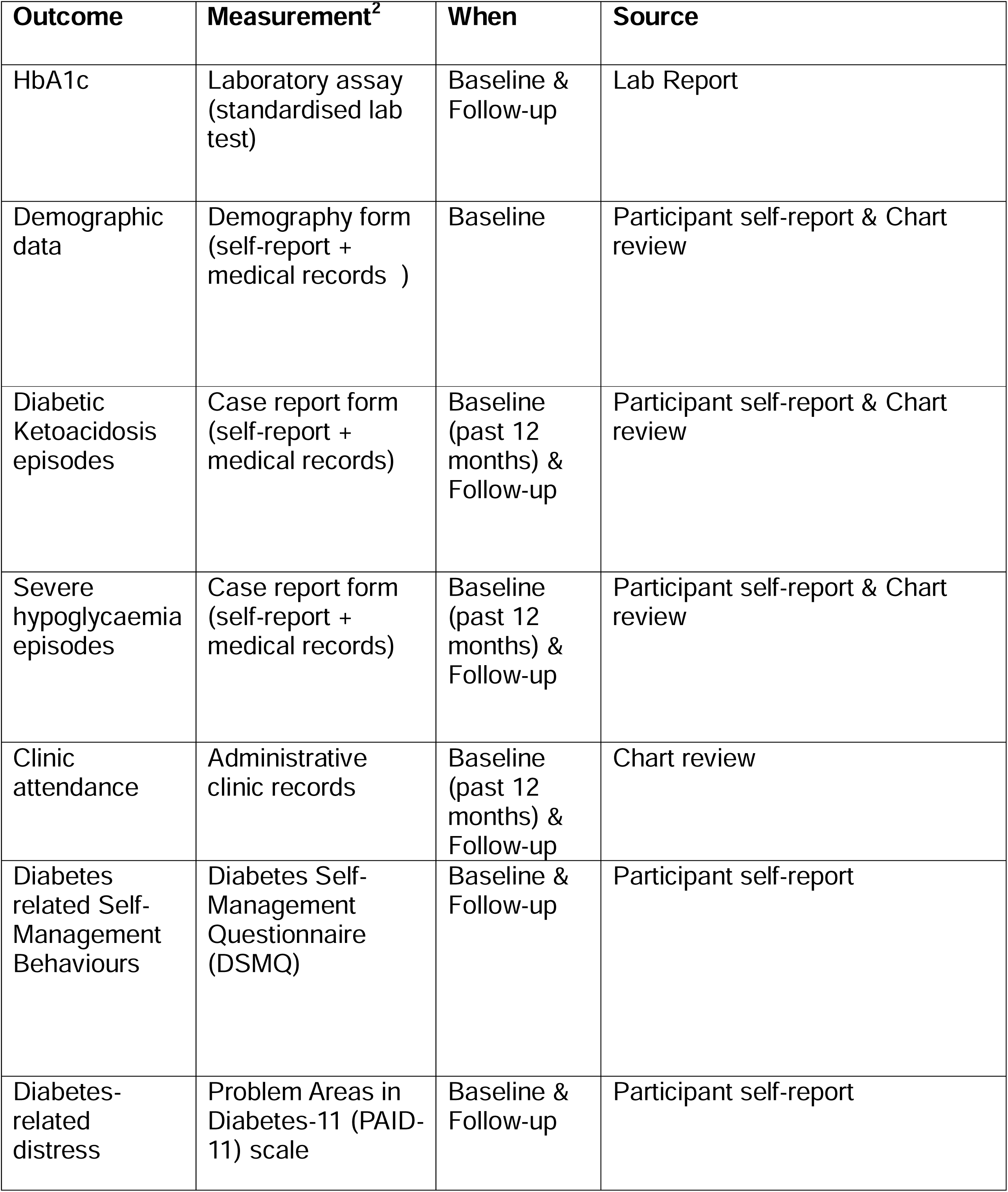
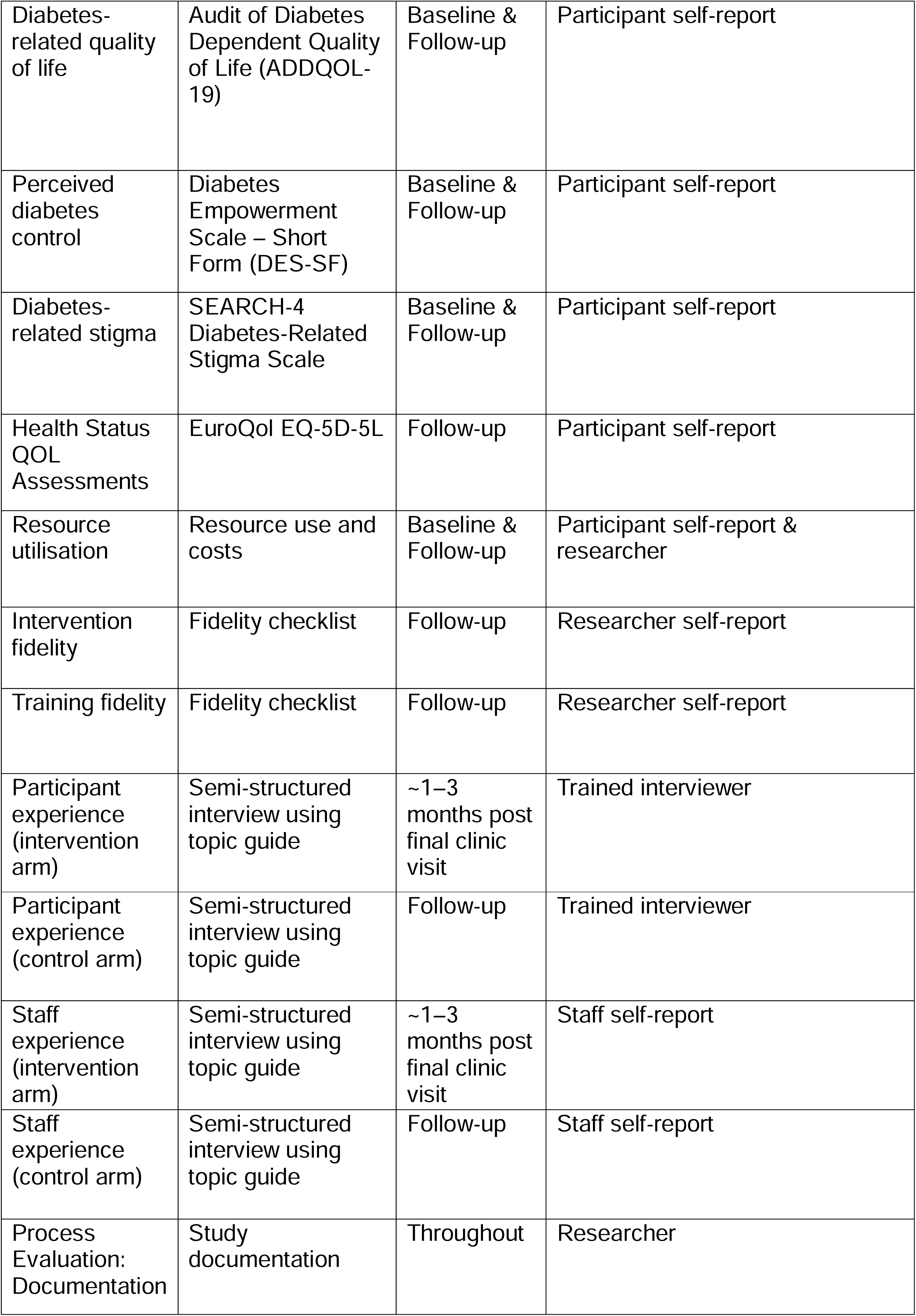
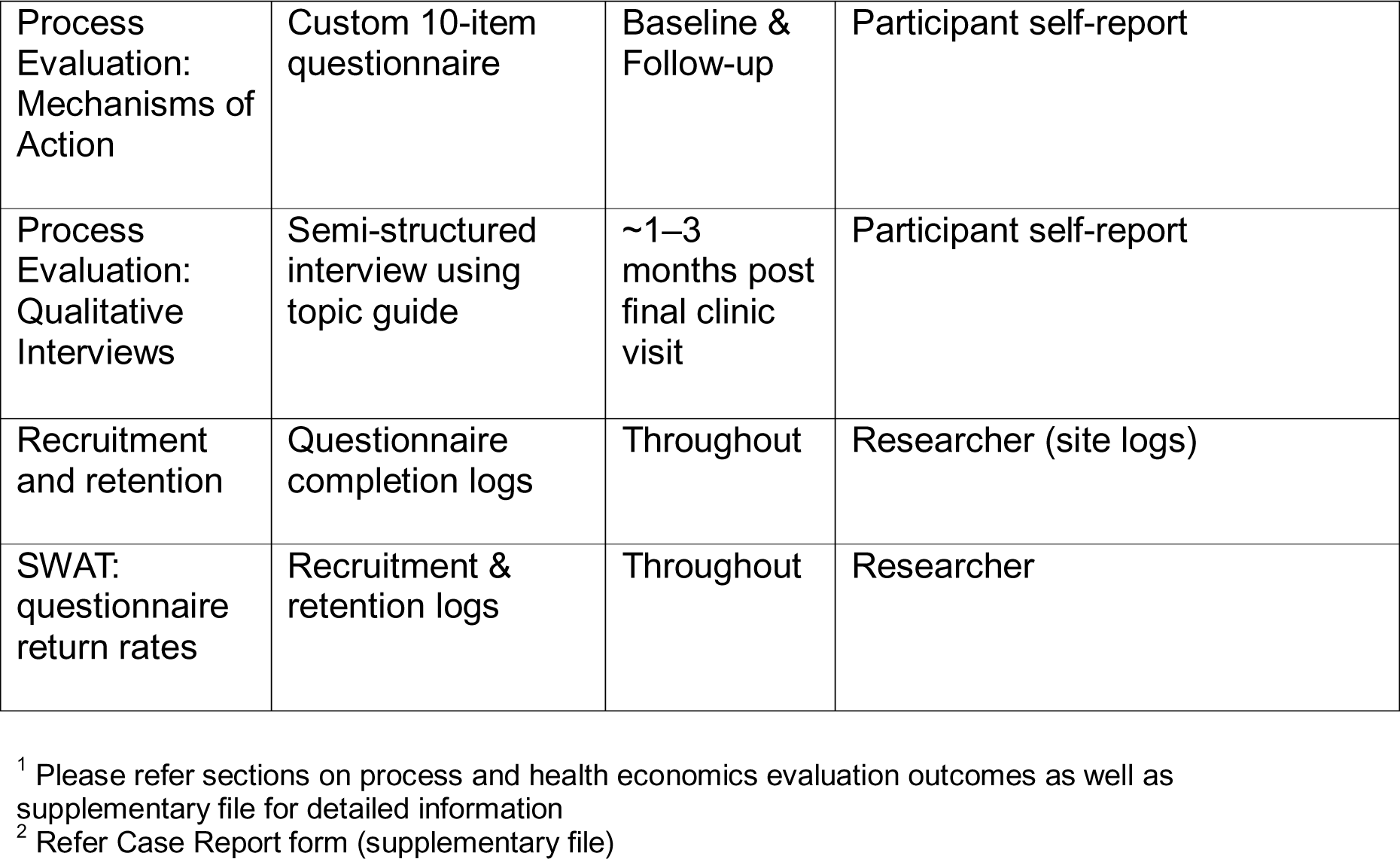
Description of Outcomes^1^.

### Sample Size Calculation

We aim to recruit 348 participants from 12 diabetes centres (6 in each arm) to ensure that at least 250 young adults complete the study. The dropout rate is based on the pilot study, where we observed a loss to follow-up of one in five centres and 15% of randomised participants. The sample size calculation assumes an intracluster correlation coefficient (ICC) of 0.0115 based on results obtained in the pilot study. If the observed ICC differs substantially, we will recalculate the required sample size. This sample size is powered to detect a clinically significant difference in the primary outcome, HbA1c levels. Assuming a standard deviation of 10 mmol/mol in HbA1c levels, this sample size will provide 90% power to detect a mean difference of 5 mmol/mol in HbA1c levels between the intervention and control groups at a significance level of 5%. The sample size calculation was guided by the revised minimum clinically important difference (MCID) of 5 mmol/mol for HbA1c, based on expert discussions and literature review, particularly insights from the DAFNEplus study^33^, nature of the intervention and the anticipated variability in clinical practice.

### Data analysis plan

We will perform all final analyses identified in the Statistical Analysis Plan after the last patient has completed the study. The primary outcome will be the difference between intervention and control groups in mean change in HbA1c between baseline and 12 months follow-up, analysed at the individual participant level, accounting for clustering. We will use linear mixed models to incorporate the homogeneity of individuals due to cluster membership, utilising a random intercepts model. We will make an adjustment for baseline HbA1c by including baseline HbA1c as a covariate when modelling HbA1c at 12 months and by modelling the change from baseline (i.e., post minus pre values for HbA1c) as the primary response. Additionally, we will include the baseline insulin delivery method and the stratification variable hospital model as adjustment variables. We will address any chance imbalances between treatment groups in key covariates, such as age and history of severe hypoglycaemia in the past year, by including these variables as adjustments in the statistical models. We will examine differences between adjusted and unadjusted estimates to assess the impact of these imbalances.

Analysis will follow intention-to-treat principles, with all randomised participants analysed in their assigned groups regardless of intervention receipt. A per-protocol sensitivity analysis will include only participants without major protocol deviations who attended at least two of the three scheduled intervention visits, with all randomised participants analysed in their assigned groups, regardless of intervention receipt. A per-protocol sensitivity analysis will include only participants who attended at least 2 of 3 scheduled intervention visits

A detailed statistical analysis plan is described in the supplementary files.

### Secondary study aims

#### Health economic evaluation

We will incorporate a health economic evaluation to determine the cost-effectiveness of the intervention compared with standard care. Building on the methodological approaches developed in the pilot study, we will undertake an incremental trial-based cost-utility analysis from a healthcare perspective. Participants will complete a health economics questionnaire at baseline and at 12 months, capturing data on healthcare resource utilisation and preference-based outcomes. Unit costs will be applied to value resource activity and generate total cost variables, and data collected using the EuroQol EQ-5D-5L instrument will be used to generate Quality-Adjusted Life Years (QALYs). The economic evaluation will be informed by national guidelines for the conduct of health technology assessment, incorporating deterministic and probabilistic sensitivity analyses, and supplementary subgroup and modelling analyses.

#### Process Evaluation and Qualitative Evaluation

We will embed a detailed process evaluation within the trial using a convergent parallel mixed-methods design. We will publish a separate process evaluation protocol that explains how we are assessing fidelity, how the intervention is being delivered in real-world settings, and what factors might influence that delivery. The protocol will cover how we are collecting and analysing this information, including researcher logs, fidelity checklists, and interviews with participants and staff regarding their experiences with the intervention. It will also describe the training and support that will be provided to help sites deliver the intervention as intended and to build consistency across clinics.

This evaluation aims to assess the fidelity of intervention delivery, explore hypothesised mechanisms of action, and understand how the intervention is implemented across different clinic contexts. We will administer a bespoke 10-item mechanisms of action questionnaire to all young adult participants online at both time points to assess the intervention’s impact on key theoretical constructs underpinning the logic model. Additionally, we will use fidelity checklists to evaluate the consistency of training and delivery across sites, which will be completed by both support workers and participants following clinic visits. The process evaluation will also track study metrics such as clinic attendance and support worker contact frequency, allowing for assessment of intervention reach and engagement.

To complement these quantitative findings, we will conduct a rich qualitative study following the intervention. We will invite a subset of 54 young adult participants, 21 staff members (from both intervention and control groups), and support workers for semi-structured interviews, scheduled within three months after the follow-up point. These interviews will explore experiences with the D1 Now intervention and provide insight into the mechanisms of action and contextual factors that affect implementation.

The qualitative insights will be triangulated with quantitative findings to enrich the understanding of the intervention’s impact, support the interpretation of outcome data, and guide the refinement of the intervention for future implementation.

## Conclusions

Young adults with T1D face disproportionately high rates of acute complications and disengagement from structured diabetes care during a period marked by substantial life transitions. The D1 Now intervention directly responds to this reality by embedding structured behavioural supports within routine clinical pathways. By integrating a support worker, which is hypothesised to enhance relational continuity with healthcare professionals and an Agenda-setting tool, the intervention seeks to address not just clinical targets but the relational and contextual factors that influence engagement.

This definitive cluster randomised trial builds directly on the learning from a prior pilot study, which tested intervention feasibility across four clinics. The current trial extends its reach to a national level and incorporates refinements based on pilot findings, including improved recruitment strategies and strengthened implementation procedures. A mixed-methods process evaluation and health economic analysis will provide insights into how the intervention works in routine care, its cost-effectiveness, and its practical implications for national rollout.

The trial’s design reflects the complexity of young adult care and prioritises real-world applicability by embedding the intervention into existing clinical structures. If effective, the D1 Now intervention has the potential to offer a scalable model of care tailored to the needs of young adults with T1D in Ireland and beyond.

## Data Availability Statement

The dataset supporting the conclusions of this article is included within the article. Additional materials and data supporting this project are available in the supplementary files and via the Open Science Framework (OSF) repository https://osf.io/qyw3p/. A detailed version of the full protocol, along with supplementary study materials, will be made publicly available on the OSF repository once ethics approval has been obtained from all participating committees.

## Protocol amendments

Any modification to the study protocol which may impact the conduct of the study or the potential safety of or benefits to participants, including changes to the study objectives, study design, sample size or study procedures, will require a formal amendment to the protocol. Such amendments will require approval from all relevant Ethics Committees.

## Funding

The study is funded by the Health Research Board (HRB), Ireland (DIFA-2023-018). The HRB had no role in the design of the intervention or data collection tools and will not influence the conduct, analysis, or reporting of trial results. All decisions relating to design, analysis, and dissemination rest with the research team and oversight committees.

## Corresponding authors

Correspondence to Aswathi Surendran.

## Ethics declarations

### Ethics approval and consent to participate

As a multi-site trial, the study sought approval from multiple ethics committees^2^, and all study participants will give consent to participate in the proposed study.

### Consent for publication

Not applicable to the protocol. Participants will provide written informed consent for the use of their anonymised data and quotes in resultant publications.

### Funder

The D1 Now study is funded by the Health Research Board (DIFA-2023-018). The Health Research Board has no role in the study design, data collection or analysis, or decision to publish or prepare the manuscript.

Dr Aswathi Surendran was supported by the Health Research Board (Ireland) (Grant number TMRN-2021-001) through the HRB – Trials Methodology Research Network (HRB–TMRN).

### Sponsor

The University of Galway will serve as the sponsor for the study, with the Chief Investigator (CI) assuming primary responsibility for overseeing the trial. The sponsor will not be involved in data collection, analysis, data interpretation, report writing, or the decision to publish the report.

### Insurance and Indemnity

The Sponsor for this trial is the University of Galway (formerly NUI Galway), the Chief Investigator’s employing institution. As such, the CI, under the sponsorship of the host University, bears ultimate responsibility for the study and compliance with the regulations, principles and standards of good clinical practice that govern clinical research. The Sponsor assumes responsibility for initiating, managing, and reporting the clinical trial and its interventions. As the trial Sponsor, University of Galway is responsible for ensuring that appropriate insurance or indemnity is in place as per the HRB general terms and conditions to cover liabilities which may arise in relation to the design, management and conduct of the trial.

## Abbreviations

ADDQOL: Audit of Diabetes-Dependent Quality of Life
CI: Chief Investigator
CONSORT: Consolidated Standards of Reporting Trials
CRF: Case Report Form
CMG: Core Study Management Group
DAFNE: Dose Adjustment for Normal Eating
DDS: Diabetes Distress Scale
DES-SF: Diabetes Empowerment Scale-Short Form
DKA: Diabetic Ketoacidosis
DPIA: Data Protection Impact Assessment
DSMQ-R: Diabetes Self-Management Questionnaire - Revised
HbA1c: Haemoglobin A1c
HIQA: Health Information and Quality Authority
HRB: Health Research Board
HRB-CRFG: Health Research Board-Clinical Research Facility Galway
ICF: Informed Consent Form
HSE: Health Service Executive
HSC: Health and Social Care
ITSDMC: Independent Trial Steering and Data Monitoring Committee
PAID-11: Problem Areas in Diabetes -11
PI: Principal Investigator
PPI: Patient and Public Involvement
PIS/PIL: Participant Information Sheet/Leaflet
QALs: Quality-adjusted Life Years
RCT: Randomised Control Trial
SPIRIT: Standard Protocol Items: Recommendations for Interventional Trials
SOP: Standard Operating Procedure
SWAT: Study Within a Trial
T1-DDS: Type 1 Diabetes Distress Scale
TMF: Trial Master File
TMG: Trial Management Group
YAP: Young Adult Panel

## Data Availability

All data produced in the present study are available upon reasonable request to the authors

https://osf.io/qyw3p/

## Acknowledgements

We would like to express our sincere gratitude to all those who have contributed to the development and delivery of the D1 Now trial. We thank the Health Research Board (HRB) for funding this research and for their continued support. We are grateful to the Research Ethics Committees for reviewing and approving the study, which enabled its ethical conduct across all participating sites.

We extend our thanks to the HRB Diabetes Collaborative Clinical Trial Network for their guidance in supporting multi-site implementation, and to the Clinical Research Facility Galway for their expertise in quality oversight, data management and operational coordination. We deeply appreciate the ongoing insights and contributions from the D1 Now Young Adult Panel (YAP) Mentors, who have helped shape the study to be meaningful and relevant to young adults living with type 1 diabetes.

We also thank the Trial Management Group for their oversight and dedication, and the Independent Trial Steering and Data Management Committee (ITSDMC) for their support in establishing secure and effective oversight. Finally, we acknowledge the Principal Investigators and site teams at each participating centre for their continuous support, input, and commitment throughout the trial preparation phase.

We are grateful to Prof Dympna Casey for providing qualitative expertise in the early phase of protocol development.

## Footnotes

1 https://www.gov.ie/en/department-of-health/collections/type-1-diabetes-mellitus-in-adults-version-2/

2 REC approval status is regularly updated in the OSF page: https://osf.io/3uj6k/

## Notes

### Competing Interest Statement

The authors have declared no competing interest.

### Clinical Trial

ISRCTN28944606

### Author Declarations

This study will be conducted, analysed, and reported according to the Consolidated Standards of Reporting Trials (CONSORT) 2025 statement extension for cluster RCTs.27 Ethical approval will be sought from the relevant Research ethics committees. Ethics Committee of Galway University Hospital gave ethical approval for this work. Consent for publication Not applicable to the protocol. Participants will provide written informed consent for the use of their anonymised data and quotes in resultant publications.

## Reference

1. Garvey KC, Markowitz JT, Laffel LMB. Transition to Adult Care for Youth with Type 1 Diabetes. Curr Diab Rep. 2012;12(5):533–541. doi:10.1007/s11892-012-0311-6

2. Sparud-Lundin C, Öhrn I, Danielson E. Redefining relationships and identity in young adults with type 1 diabetes. J Adv Nurs. 2010;66(1):128–138. doi:10.1111/j.1365-2648.2009.05166.x

3. Morrissey EC, Casey B, Dinneen SF, Lowry M, Byrne M. Diabetes Distress in Adolescents and Young Adults Living With Type 1 Diabetes. Can J Diabetes. 2020;44(6):537–540. doi:10.1016/j.jcjd.2020.03.001

4. Morrissey EC, Dinneen SF, Lowry M, de Koning EJ, Kunneman M. Reimagining care for young adults living with typelJ1 diabetes. J Diabetes Investig. 2022;13(8):1294–1299. doi:10.1111/jdi.13824

5. Laursen MG, Rahbæk MØ, Jensen SD, Prætorius T. Experiences of young people living with type 1 diabetes in transition to adulthood: The importance of care provider familiarity and support. Scand J Caring Sci. 2024;38(1):126–135. doi:10.1111/scs.13214

6. O’Hara MC, Hynes L, O’donnell M, et al. A systematic review of interventions to improve outcomes for young adults with Type 1 diabetes. Diabet Med. 2017;34(6):753–769.

7. Casey R, O’Hara MC, Cunningham A, et al. Young adult type 1 diabetes care in the West of Ireland: an audit of hospital practice. QJM Int J Med. 2014;107(11):903–908.

8. Monaghan M, Helgeson V, Wiebe D. Type 1 diabetes in young adulthood. Curr Diabetes Rev. 2015;11(4):239–250. doi:10.2174/1573399811666150421114957

9. Bryden KS, Dunger DB, Mayou RA, Peveler RC, Neil HAW. Poor prognosis of young adults with type 1 diabetes: a longitudinal study. Diabetes Care. 2003;26(4):1052–1057.

10. James S, Perry L, Lowe J, et al. Suboptimal glycemic control in adolescents and young adults with type 1 diabetes from 2011 to 2020 across Australia and New Zealand: Data from the Australasian Diabetes Data Network registry. Pediatr Diabetes. 2022;23(6):736–741.

11. Hislop AL, Fegan PG, Schlaeppi MJ, Duck M, Yeap BB. Prevalence and associations of psychological distress in young adults with Type 1 diabetes. Diabet Med J Br Diabet Assoc. 2008;25(1):91–96. doi:10.1111/j.1464-5491.2007.02310.x

12. Hanna KM, Woodward J. The Transition From Pediatric to Adult Diabetes Care Services. Clin Nurse Spec. 2013;27(3):132. doi:10.1097/NUR.0b013e31828c8372

13. Hynes L, Byrne M, Dinneen SF, McGuire BE, O’Donnell M, Mc Sharry J. Barriers and facilitators associated with attendance at hospital diabetes clinics among young adults (15–30 years) with type 1 diabetes mellitus: a systematic review. Pediatr Diabetes. 2016;17(7):509–518. doi:10.1111/pedi.12198

14. Walsh DM, Hynes L, O’Hara MC, Mc Sharry J, Dinneen SF, Byrne M. Embedding a user-centred approach in the development of complex behaviour change intervention to improve outcomes for young adults living with type 1 diabetes: The D1 Now Study. HRB Open Res. 2018;1. doi:10.12688/hrbopenres.12803.2

15. Casey B, Byrne M, Casey D, et al. Improving outcomes among young adults with type 1 diabetes: the D1 Now Randomised Pilot Study Protocol. Diabet Med. 2020;37(9):1590–1604.

16. Morrissey EC, Casey B, Hynes L, Dinneen SF, Byrne M, The D1 Now Young Adult Panel. Supporting self-management and clinic attendance in young adults with type 1 diabetes: development of the D1 Now intervention. Pilot Feasibility Stud. 2021;7(1):186. doi:10.1186/s40814-021-00922-z

17. Hynes L, Byrne M, Casey D, Dinneen SF, O’Hara MC. ‘It makes a difference, coming here’: A qualitative exploration of clinic attendance among young adults with type 1 diabetes. Br J Health Psychol. 2015;20(4):842–858. doi:10.1111/bjhp.12145

18. O’Hara MC, Hynes L, O’Donnell M, et al. Strength in Numbers: an international consensus conference to develop a novel approach to care delivery for young adults with type 1 diabetes, the D1 Now Study. Res Involv Engagem. 2017;3(1):25. doi:10.1186/s40900-017-0076-9

19. McCarthy EM, Dinneen SF, Byrne M, Morrissey EC, Casey D. Perceptions and experiences of young adults and their healthcare team of the D1 Now type 1 diabetes intervention. PLOS ONE. 2025;20(2):e0316345. doi:10.1371/journal.pone.0316345

20. Morrissey EC, Byrne M, Casey B, et al. Improving outcomes among young adults with type 1 diabetes: the D1 Now pilot cluster randomised controlled trial. Pilot Feasibility Stud. 2022;8(1):56.

21. Choudhary P, Edwards F, Patel NH, Spratling L, Sturt J. Type 1 Consultation (T1C) Tool. South London Health Innovation Network; 2016. https://healthinnovationnetwork.com/wp-content/uploads/2021/08/TC1-toolkit-final.pdf

22. McCarthy EM, Dinneen SF, Byrne M, Morrissey EC, Casey D. Perceptions and experiences of young adults and their healthcare team of the D1 Now type 1 diabetes intervention. PLOS ONE. 2025;20(2):e0316345. doi:10.1371/journal.pone.0316345

23. Lowry M, Morrissey EC, Dinneen SF. Piloting an Intervention to Improve Outcomes in Young Adults Living With Type 1 Diabetes: The Experience of the D1 Now Support Worker. Front Clin Diabetes Healthc. 2021;2:799589. doi:10.3389/fcdhc.2021.799589

24. O’Hara MC, Cunningham Á, Keighron C, et al. Formation of a type 1 diabetes young adult patient and public involvement panel to develop a health behaviour change intervention: the D1 Now study. Res Involv Engagem. 2017;3(1):21. doi:10.1186/s40900-017-0068-9

25. Eimear Morrissey, Blathin Casey, Rachael Power, D1 Now Young Adult Panel. SWATlJ133: Branded Gift and Letter from PPI Group to Enhance Questionnaire Response Rate in a Randomised Trial. Northern Ireland Network for Trials Methodology Research, Queen’s University Belfast https://www.qub.ac.uk/sites/TheNorthernIrelandNetworkforTrialsMethodologyResearch/FileStore/Filetoupload,998532,en.pdf

26. Young Adult Panel Recruitment – D1 Now. Accessed July 9, 2025. https://d1now.ie/pages/young-adult-panel-recruitment

27. Hopewell S, Chan AW, Collins GS, et al. CONSORT 2025 statement: updated guideline for reporting randomised trials. BMJ. 2025;389:e081123. doi:10.1136/bmj-2024-081123

28. SPIRIT 2025 statement: updated guideline for protocols of randomised trials | The BMJ. Accessed July 28, 2025. https://www.bmj.com/content/389/bmj-2024-081477

29. Fisher L, Glasgow RE, Mullan JT, Skaff MM, Polonsky WH. Development of a brief diabetes distress screening instrument. Ann Fam Med. 2008;6(3):246–252. doi:10.1370/afm.842

30. Polonsky WH, Fisher L, Earles J, et al. Assessing Psychosocial Distress in Diabetes: Development of the Diabetes Distress Scale. Diabetes Care. 2005;28(3):626–631. doi:10.2337/diacare.28.3.626

31. Nygaard M, Willaing I, Joensen LE, et al. A Short-Form Measure of Diabetes Distress Among Adults With Type 1 Diabetes for Use in Clinical Practice: Development and Validation of the T1-DDS-7. Diabetes Care. 2023;46(9):1619–1625. doi:10.2337/dc23-0460

32. Byrne M, O’Connell A, Egan AM, et al. A core outcomes set for clinical trials of interventions for young adults with type 1 diabetes: an international, multi-perspective Delphi consensus study. Trials. 2017;18(1):602. doi:10.1186/s13063-017-2364-y

33. Heller S, Lawton J, Amiel S, et al. Improving management of type 1 diabetes in the UK: the Dose Adjustment For Normal Eating (DAFNE) programme as a research test-bed. A mixed-method analysis of the barriers to and facilitators of successful diabetes self-management, a health economic analysis, a cluster randomised controlled trial of different models of delivery of an educational intervention and the potential of insulin pumps and additional educator input to improve outcomes. Programme Grants Appl Res. 2014;2(5).

