## Appendix table for "Evaluating the effectiveness of the D1 Now intervention to improve outcomes among young adults with type 1 diabetes: Protocol for a cluster randomised controlled trial"

### Supplementary File

#### Table S1 Detailed Data Collection Summary

| **Data Collected** | **Measurement** | **Measurement Tool** | | **Time Points** | **Collected by** | **Storage Location** | | **Purpose** | **Sub-study** |
| --- | --- | --- | --- | --- | --- | --- | --- | --- | --- |
| **Haemoglobin A1c (HbA1c) levels** | Standardised Laboratory assays | Haemoglobin A1c laboratory test | | Baseline, Follow-up | Laboratory technician | Central Laboratory, CRF Galway Secure Database | | To measure the primary outcome of diabetes control effectiveness. | Main Study |
| **Demographic data** | Self-reported and case report form | Demography form | | Baseline | Onsite Study coordinator | CRF Galway Secure Database | | To capture participant's baseline demographic details. To stratify analyses by demographic factors (if required) and assess representativeness. | Main Study |
| **Episodes of Diabetic Ketoacidosis (DKA)** | Self-report and medical records | Case report form | | Baseline (past 12 months) & Follow-up | Onsite Study Coordinator | CRF Galway Secure Database | | To track and analyse critical DKA episodes. | Main Study |
| **Episodes of Severe Hypoglycemia** | Self-report and medical records | Case report form | | Baseline (past 12 months) & Follow-up | Onsite Study Coordinator | CRF Galway Secure Database | | To document occurrences of severe hypoglycemia. | Main Study |
| **Clinic Attendance** | Clinic administration records | Administrative records | | Baseline (past 12 months) & Follow-up | Clinic administration | CRF Galway Secure Database | | To evaluate changes in the participant engagement with clinical care due to the intervention. | Main Study |
| **Diabetes related Self-Management Behaviours^[1]^** | Self-report measures | Diabetes Self-Management Questionnaire (DSMQ) | | Baseline and Follow-up | Onsite Study coordinator | CRF Galway Secure Database | | To evaluate changes in self-management behaviours due to the intervention. | Main Study |
| **Diabetes-Related Distress^[2]^** | Self-report measures | Problem Areas in Diabetes-11 (PAID-11) scale | | Baseline and Follow-up | Onsite Study coordinator | CRF Galway Secure Database | | To assess psychological impacts and intervention effectiveness on distress. | Main Study |
| **Diabetes-Related Quality of Life^[3]^** | Self-report measures | Audit of Diabetes Dependent Quality of Life (ADDQOL-19) | | Baseline and Follow-up | Onsite Study coordinator | CRF Galway Secure Database | | To determine the intervention's effects on life quality. | Main Study |
| **Perceived Level of Control Over Diabetes^[4]^** | Self-report measures | Diabetes Empowerment Scale–Short Form (DES-SF) | | Baseline and Follow-up | Onsite Study coordinator | CRF Galway Secure Database | | To measure intervention's effects on perceived control over diabetes. | Main Study |
| **Diabetes-related stigma^[5]^** | Self-report measures | SEARCH 4 Diabetes-Related Stigma Survey Scale | Baseline and Follow-up at 12 months | | Onsite Study coordinator | | CRF Galway Secure Database | To assess the frequency of stigmatising experiences among young adults with diabetes. | Main Study |
| **Participant experiences - Intervention arm** | Semi-structured interviews | Interview guide | | Follow-up | Trained interviewers | University of Galway One drive Storage | | To gather insights on participant experiences with the intervention. | Process evaluation and qualitative study |
| **Participant experiences - Control arm** | Semi-structured interviews | Interview guide | | Follow-up | Trained interviewers | University of Galway One drive Storage | | To understand participant experiences under standard care. | Process evaluation and qualitative study |
| **Healthcare staff experiences - Intervention arm** | Semi-structured interviews | Interview guide | | Follow-up | Trained interviewers | University of Galway One drive Storage | | To capture healthcare staff experiences with the intervention. | Process evaluation and qualitative study |
| **Healthcare staff experiences - Control arm** | Semi-structured interviews | Interview guide | | Follow-up | Trained interviewers | University of Galway One drive Storage | | To understand healthcare staff experiences with standard care. | Process evaluation and qualitative study |
| **Resource utilisation** | Self-report and research report | Resource utilisation, quality-adjusted life years (QALYs) | | Baseline and Follow-up | Onsite Study coordinator | University of Galway One drive Storage | | To assess cost-effectiveness of the intervention for health economic evaluation. | Economic Analysis |
| **Health Status QOL Assessments^[6]^** | Self-report measures | EuroQol EQ-5D-5L instrument | | Follow-up | Onsite Study coordinator | CRF Galway Secure Database | | To evaluate the general health status and quality of life for health economic evaluation. | Economic Analysis |
| **Adherence to intervention protocols** | Self-report and research report | Intervention fidelity checklist | | Follow-up | Onsite Study coordinator | CRF Galway Secure Database | | To monitor fidelity in implementing the intervention. | Process evaluation and qualitative study |
| **Recruitment and Retention Rate** | Recruitment report and research report | Questionnaire completion | | Throughout Trial | Onsite Study coordinator | CRF Galway Secure Database | | To track and record recruitment efforts and demographics. | Process evaluation and qualitative study |
| **Process Evaluation: Documentation** | Recruitment, approval, demographics | Study documentation | | Ongoing | Study team | University of Galway One drive Storage and secured university lockers | | To ensure proper reporting and documentation of trial processes. | Process evaluation and qualitative study |
| **Process Evaluation: Fidelity Checks (Training)** | Fidelity to training protocols | Fidelity checklist | | Ongoing | Study team and clinic staff | CRF Galway Secure Database | | To monitor, document and evaluate the implementation of training components. | Process evaluation and qualitative study |
| **Process Evaluation: Fidelity Checks (Intervention)** | Fidelity to intervention protocols | Fidelity checklist | | Ongoing | Support Workers and Young Adult participants | CRF Galway Secure Database | | To monitor, document and evaluate the implementation of intervention components. | Process evaluation and qualitative study |
| **Process Evaluation: Mechanisms of Action** | Questionnaire on mechanisms of action | Custom-designed 10-item questionnaire | | Baseline and Follow-up | Study team | CRF Galway Secure Database | | To test the proposed mechanisms of impact within the intervention logic model. | Process evaluation and qualitative study |
| **Process Evaluation: Qualitative Interviews** | Perceptions and experiences | Semi-structured interview guides for young adults with T1D1, and healthcare professionals in the intervention and control groups and support workers | | Follow-up | Trained interviewers | University of Galway One drive Storage | | To explore the perceptions and experiences of participants and staff regarding the intervention and standard care. | Process evaluation and qualitative study |
| **Study within a Trial (SWAT)** | Number and proportion of participants who complete the final study questionnaire | Recruitment and retention log | | Throughout Trial | Randomised intervention by PPI group | CRF Galway Secure Database and University of Galway One drive Storage | | To measure the effectiveness of the SWAT intervention in improving questionnaire response rates. | SWAT |

#### Figure S1 Study Flow

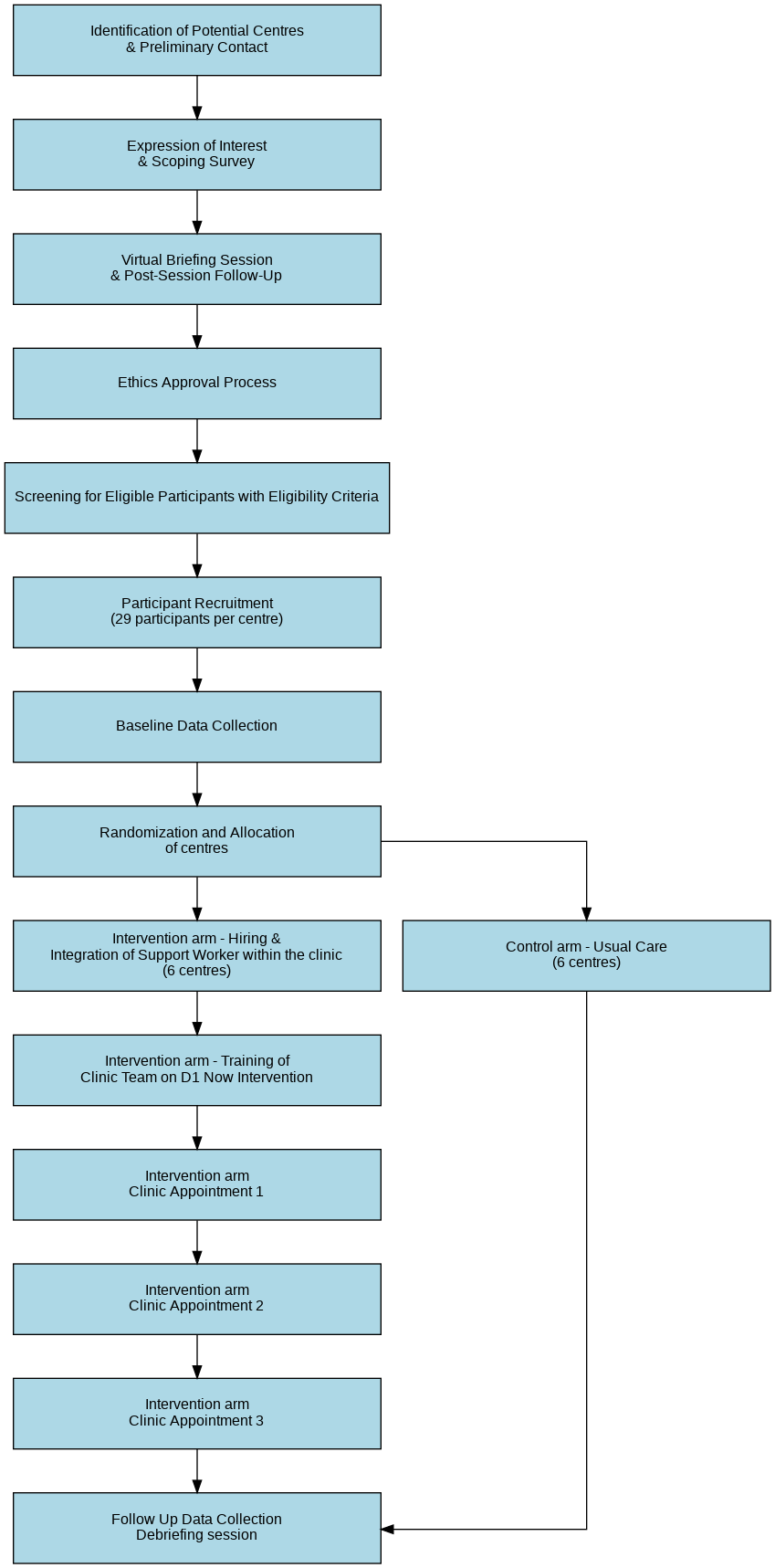

#### Figure S2 Agenda setting Tool

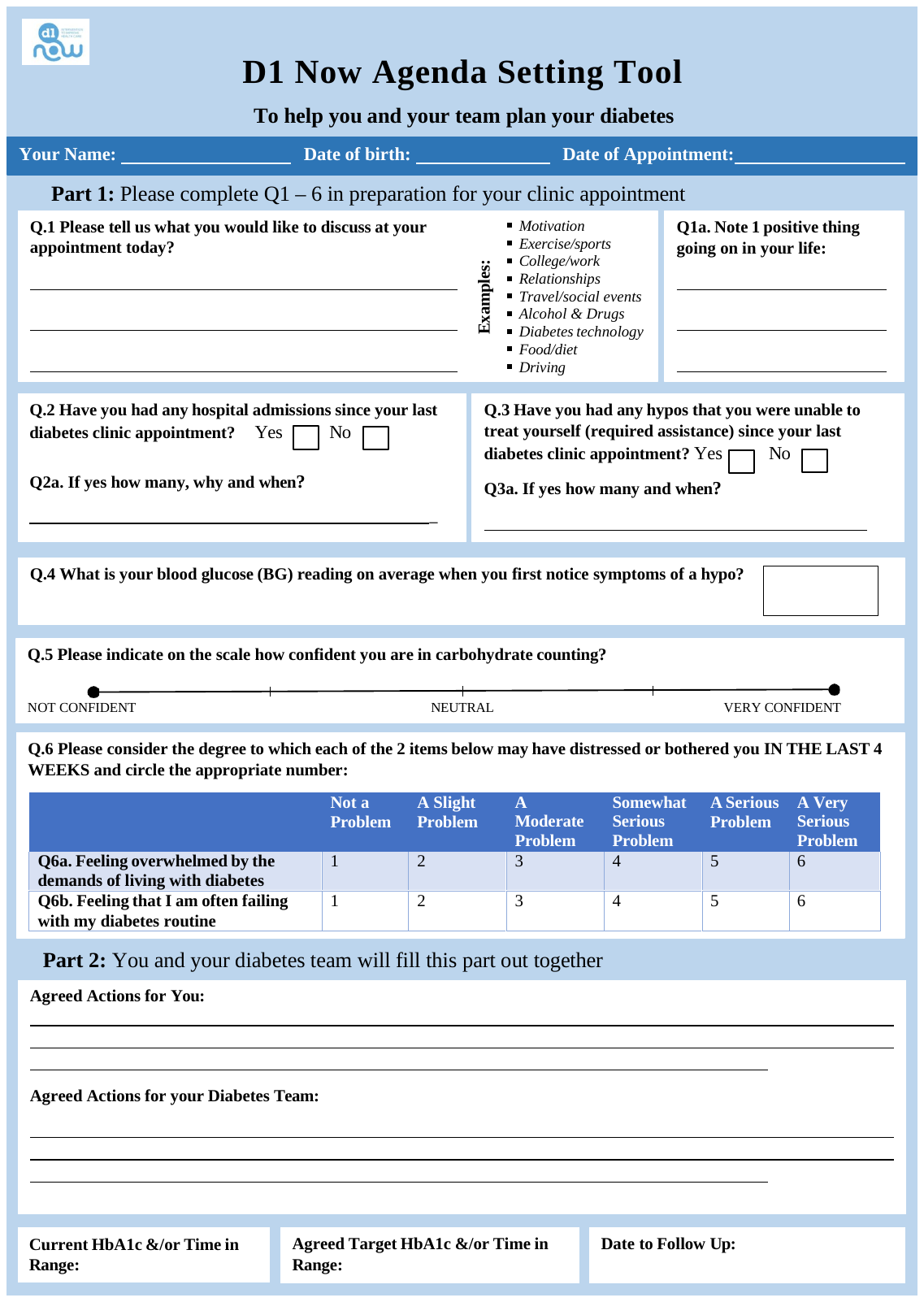

#### Table S2 Schedule of events

Table 2: Schedule of events

|  | **STUDY PERIOD** | | | | | | | |
| --- | --- | --- | --- | --- | --- | --- | --- | --- |
|  | **Enrolment** | **Allocation** | **Post-allocation^[7]^** | | | | | **Close-out** |
| **TIMEPOINT**** | ***-t_1_*** | ***t_1_*** | ***t_0_*** | ***t_2_*** | ***t_3_*** | ***t_4_*** | ***t_5_*** | ***t_x_*** |
| **ENROLMENT:** |  |  |  |  |  |  |  |  |
| ***Eligibility screen*** | X |  |  |  |  |  |  |  |
| ***Informed consent*** | X |  |  |  |  |  |  |  |
| ***Randomisation*** |  | X |  |  |  |  |  |  |
| ***Allocation*** |  | X |  |  |  |  |  |  |
| ***Patient demography*** |  |  | X |  |  |  |  |  |
| **INTERVENTION:** |  |  |  |  |  |  |  |  |
| ***Agenda Setting tool*** |  |  |  | X | X | X |  |  |
| ***Support Worker*** |  |  |  | X | X | X |  |  |
| **ASSESSMENTS:** |  |  |  |  |  |  |  |  |
| ***Patient medical history*** |  |  | X |  |  |  | X |  |
| ***Psychosocial and diabetes management questionnaires*** |  |  | X |  |  |  | X |  |
| ***Laboratory testing - HbA1c*** |  |  | X |  |  | X |  |  |
| ***Clinical outcomes*** |  |  | X |  |  |  | X |  |
| ***Process evaluation questionnaires*** |  |  | X |  |  |  | X |  |
| ***Health economics questionnaires*** |  |  | X |  |  |  | X |  |
| ***Clinical Attendance Report*** |  |  |  | X | X | X |  |  |
| ***Concomitant Medication Report*** |  |  |  | X | X | X |  |  |
| ***Adverse events*** |  |  |  | X | X | X |  |  |
| ***Fidelity questionnaires*** |  |  |  | X | X | X |  |  |
| ***Follow up interview*** |  |  |  |  |  |  | X |  |
| ***Complete end of study form*** |  |  |  |  |  |  |  | X |

#### Section S1 Intervention Manual

<https://osf.io/bpuzf/?view_only=80f9662c2d0948f6b755d4363803d675>

#### Section S2 Case Report Form

<https://osf.io/bpuzf/?view_only=80f9662c2d0948f6b755d4363803d675>

#### Section S3 Roles and Responsibilities

Study Oversight Committees

The Chief Investigator (CI) holds overall responsibility for managing the D1 Now study. Oversight of the day-to-day operations and coordination of the multi-disciplinary trial team is the responsibility of the Programme Manager. The study is co-ordinated from the Health Research Board-Clinical Research Facility Galway (HRB-CRFG) under a robust quality management system that complies with relevant standards and guidelines.

Additionally, specific oversight committees are formed for the D1 Now study, including a Core Study Management Group (CMG), Trial Management Group (TMG), Independent Trial Steering and Data Monitoring Committee (ITSDMC) and Scientific Advisory Board (SAB).

Core Study Management Group

The Core Study Management Group (CMG) is responsible for the day-to-day management of the trial. This committee includes Prof Sean Dinneen, Prof Molly Byrne (Co-applicant), Dr Eimear Morrissey (Co-applicant) and Dr Aswathi Surendran (Programme Manager).

Trial Management Group

A Trial Management Group (TMG) is established and chaired by the Principal Investigator (PI)^[8]^. It is composed of co-applicants and collaborators. The TMG is responsible for the trial management, overall conduct, and progress of the trial. Observers may be invited to attend TMG meetings to provide expert opinions. The TMG meet face-to-face or online on a monthly/quarterly basis based on the level of activity. CMG and TMG members may also individually communicate between meeting times as needed.

Independent Trial Steering and Data Monitoring Committee

The Independent Trial Steering and Data Monitoring Committee (ITSDMC) is an independent body responsible for providing comprehensive oversight of the clinical trial on behalf of the Sponsor. A group of experienced clinicians, statisticians, researchers and trialists will act as an ITSDMC with an independent Chair and majority independent membership. Its key role is to ensure the trial is conducted in accordance with regulations and standards, monitor emerging safety and efficacy data (with the independent statistician carrying out IDMC responsibilities), make recommendations on any safety or ethical concerns, and oversee the overall IT governance to support the trial's objectives. The independent statistician will consider the need for any interim analysis, advise the ITSDMC regarding the release of data and/or information, and will consider data emerging from other related studies. The ITSDMC will meet once a year.

Data Management Team

Data Management Team, based at Clinical Research Facility Galway, manages the REDCap platform, data access controls, and data quality monitoring in collaboration with the research team.

User Involvement or any other relevant committees

Patient and Public Involvement (PPI) has been a cornerstone of the D1 Now study since its inception. The Young Adult Panel (YAP), consisting of 9 young adults living with type 1 diabetes from different regions of Ireland, serves as a co-researcher, offering lived experience expertise to guide all aspects of the study, from conceptualisation to practical implementation. The contribution of the D1Now YAP will include (but not be limited to) developing recruitment materials, designing questionnaires, developing website content, social media engagement, overseeing the Study within a Trial (SWAT) sub-study, assisting with interpretation of results and shaping dissemination strategies. For the Definitive Trial, a new cohort of YAP members will be recruited, with existing YAP members serving as mentors.

#### Section S4 Statistical Analysis Plan

<https://osf.io/bpuzf/?view_only=80f9662c2d0948f6b755d4363803d675>

#### Section S5 Monitoring, Quality Control, and Assurance

Role of Monitors

The study will involve several monitoring bodies, including the CMG and ITSDMC. These groups will oversee the trial's conduct, review interim data if needed, ensure participant safety, and maintain trial integrity.

Assurance on Good Clinical Practice and Research Governance

The study will adhere to Good Clinical Practice (GCP) guidelines and all relevant research governance frameworks. All the relevant research team members will complete the GCP certification.

Other Steps to Ensure Quality of Research

Additional quality assurance measures include regular training for research staff, implementation of standard operating procedures (SOPs), and use of validated data collection tools. The study team will adapt CRFG's approved safety oversight, risk management and monitoring associated templates, SOPs and checklists to ensure consistency and compliance across all sites. This collaborative approach will help us maintain the quality and safety of the study without compromising on operational effectiveness. Data entry will be subject to double-checking and cross-validation to ensure accuracy.

##### Safety Assessment

Given the nature of the intervention, the study is considered low risk and serious adverse events (SAEs) are expected to be minimal. However, any SAEs will be recorded and reviewed by the study team. Non-serious adverse events (AEs) will also be documented and reported as per trial safety guidelines.

##### Co-enrolment Guidelines

Throughout the trial, participants in both the intervention and control groups can receive treatment at the discretion of the treating clinical team.

Potential interactions or effects that could confound the results will be carefully monitored. Participants will be instructed to report any additional treatments or changes in their health status to the trial coordinator/support worker to ensure accurate data collection and analysis. The details will then be entered into the ‘Concomitant Medications’ form.

##### End-of-Study Definition

The study is considered complete once all data collection activities outlined in the protocol have been finalised. Participants are deemed to have completed the study upon attending all required assessments, including the baseline evaluation, intervention sessions, and follow-up assessments. The data collection phase officially concludes with the completion of the follow-up assessment, which includes online surveys and qualitative interviews.

Following the final clinical appointment, participants are given ten days to complete the quantitative survey. A 3-month period is dedicated to conducting qualitative interviews with both participants and healthcare staff to obtain detailed feedback on the intervention. During this period, the study team will actively encourage maximum participation by providing necessary support and flexibility to ensure participants complete the interviews.

However, if participants do not complete these activities within this specific timeline, despite the study team's efforts, they will be marked as lost to follow-up. The formal end of the study is marked by the locking of the database for the final analysis, which occurs after participants have completed their follow-up assessments within the pre-determined end date.

The study will be terminated early under specific conditions, including:

1. Mandate by the Research Ethics Committee (REC)
2. Mandate by the Sponsor, for instance, following recommendations from the ITSDMC, such as:

2.1 Safety concerns

2.2 Failure to recruit sufficient participants or centres as specified in the recruitment plan

3. Interruption/Termination of funding.

Upon completion or early termination of the study, all oversight committees, sponsors, and funders will be notified, and a comprehensive final report will be prepared and submitted. Participants will be informed of the study’s conclusion and provided with the study debrief sheet. Rigorous statistical analysis will be conducted on the collected data, with findings compiled into a final report for dissemination through peer-reviewed publications and conference presentations. An end-of-study dissemination event will be organised to present the findings to key stakeholders, and continued engagement with the YAP will be maintained to involve them in dissemination activities and ensure study results are disseminated to patient communities.

#### Section S6 Data management

##### Source Data

Source documents are the original records from which data for the study are first recorded and from which data are transcribed into the Case Report Form (CRF). These documents include a range of materials that capture participant data throughout the study.

For this study, source data encompasses various types of records and documents:

- **Patient records:** Data collected during routine clinical visits, including medical history, diabetes management practices, and clinical notes, will serve as source documents.
- **Laboratory Reports:** The central laboratory will provide reports on HbA1c levels, which are crucial for assessing the primary outcome. These reports will be sent directly from the laboratory to the study coordinator, who will then enter the data into the corresponding CRF.
- **Agenda Setting Tool, Study Logs and Notes:** Documentation related to the study process and clinic appointments, including appointment logs, fidelity checklists, and notes from qualitative interviews, will be maintained. These logs and notes will be recorded, uploaded, and stored securely to the CRF to ensure adherence to study procedures and improve transparency.

All source documents will be securely stored to maintain confidentiality and integrity. To ensure privacy, participants will be identified by their unique study number or code on all study-specific documents rather than by their name. The unique participant study number, allocated at the time of participant onboarding, will be used to identify and track data throughout the study. The data collected from these source documents will be transferred onto a bespoke, web-based electronic -CRF designed for the study.

##### Data Protection and Access to Data

To ensure that patient confidentiality is maintained and that the study complies with the Data Protection Act 2018, all investigators and study site staff will adhere to the core principles outlined in the Act. This involves strict guidelines for the collection, storage, processing, and disclosure of personal information.

Personal information will be collected and recorded with the utmost confidentiality. Identifying details will be replaced with code and pseudo-anonymised or depersonalised data to ensure privacy.

Data Custodianship

The D1Now CI will act as the data custodian, overseeing the management and protection of study data. The CI will ensure that all data protection procedures are followed and will be responsible for addressing any data-related issues that arise.

#### Section S7 Dissemination policy

The main findings of the trial will be submitted for publication in peer-reviewed scientific journals. Findings from the trial will also be presented at national and international conferences to ensure timely dissemination to the scientific community and healthcare practitioners. Participants will be informed of the trial results through newsletters and summary reports written in lay language. Additionally, the findings will be shared with stakeholders, including healthcare providers, policymakers, and patient advocacy groups, to inform practice and policy development in diabetes care.

Individual Clinicians must undertake not to submit any part of their individual data for publication without the prior consent of the Trial Management Group. All clinics that participated in the study will be acknowledged in the Acknowledgement section.

#### Table S3 SPIRIT Checklist

**SPIRIT 2025 checklist of items to address in a randomised trial protocol***

| **Section / Topic** | **No** | **SPIRIT 2025 checklist item description** | **Reported on page no.** |
| --- | --- | --- | --- |
| **Administrative information** | | |  |
| Title and structured summary | 1a | Title stating the trial design, population, and interventions, with identification as a protocol | 1 |
|  | 1b | Structured summary of trial design and methods, including items from the Trial Registration Data Set | 4 |
| Protocol version | 2 | Version date and identifier | 2 |
| Roles and responsibilities | 3a | Names, affiliations, and roles of protocol contributors | 1-2 |
|  | 3b | Name and contact information for the trial sponsor | 15 |
|  | 3c | Role of trial sponsor and funders in design, conduct, analysis, and reporting of trial; including any authority over these activities | 15 |
|  | 3d | Composition, roles, and responsibilities of the coordinating site, steering committee, endpoint adjudication committee, data management team, and other individuals or groups overseeing the trial, if applicable | Supplementary File |
| **Open science** | | |  |
| Trial registration | 4 | Name of trial registry, identifying number (with URL), and date of registration. If not yet registered, name of intended registry | 4 |
| Protocol and statistical analysis plan | 5 | Where the trial protocol and statistical analysis plan can be accessed | Supplementary File |
| Data sharing | 6 | Where and how the individual de-identified participant data (including data dictionary), statistical code, and any other materials will be accessible | Supplementary File |
| Funding and conflicts of interest | 7a | Sources of funding and other support (e.g., supply of drugs) | 15 |
|  | 7b | Financial and other conflicts of interest for principal investigators and steering committee members | 2 |
| Dissemination policy | 8 | Plans to communicate trial results to participants, healthcare professionals, the public, and other relevant groups (e.g., reporting in trial registry, plain language summary, publication) | Supplementary File |
| **Introduction** | | |  |
| Background and rationale | 9a | Scientific background and rationale, including summary of relevant studies (published and unpublished) examining benefits and harms for each intervention | 5-7 |
|  | 9b | Explanation for choice of comparator | 10 |
| Objectives | 10 | Specific objectives related to benefits and harms | 5-7 |
| **Methods: Patient and public involvement, trial design** | | |  |
| Patient and public involvement | 11 | Details of, or plans for, patient or public involvement in the design, conduct, and reporting of the trial | 6-7 |
| Trial design | 12 | Description of trial design including type of trial (e.g., parallel group, crossover), allocation ratio, and framework (e.g., superiority, equivalence, non-inferiority, exploratory) | 7 |
| **Methods: Participants, interventions, and outcomes** | | |  |
| Trial setting | 13 | Settings (e.g., community, hospital) and locations (e.g., countries, sites) where the trial will be conducted | 7-8 |
| Eligibility criteria | 14a | Eligibility criteria for participants | 8 |
|  | 14b | If applicable, eligibility criteria for sites and for individuals who will deliver the interventions (e.g., surgeons, physiotherapists) | 8 |
| Intervention and comparator | 15a | Intervention and comparator with sufficient details to allow replication including how, when, and by whom they will be administered. If relevant, where additional materials describing the intervention and comparator (e.g., intervention manual) can be accessed | Supplementary File |
|  | 15b | Criteria for discontinuing or modifying allocated intervention/comparator for a trial participant (e.g., drug dose change in response to harms, participant request, or improving/worsening disease) | Supplementary File |
|  | 15c | Strategies to improve adherence to intervention/comparator protocols, if applicable, and any procedures for monitoring adherence (e.g., drug tablet return, sessions attended) | Supplementary File |
|  | 15d | Concomitant care that is permitted or prohibited during the trial | Supplementary File |
| Outcomes | 16 | Primary and secondary outcomes, including the specific measurement variable (e.g., systolic blood pressure), analysis metric (e.g., change from baseline, final value, time to event), method of aggregation (e.g., median, proportion), and time point for each outcome | 10-11 |
| Harms | 17 | How harms are defined and will be assessed (e.g., systematically, non-systematically) | Supplementary File |
| Participant timeline | 18 | Time schedule of enrollment, interventions (including any run-ins and washouts), assessments, and visits for participants. A schematic diagram is highly recommended (see Figure) | Supplementary File |
| Sample size | 19 | How sample size was determined, including all assumptions supporting the sample size calculation | 11 |
| Recruitment | 20 | Strategies for achieving adequate participant enrollment to reach target sample size | 8-9 |
| **Methods: Assignment of interventions** | | |  |
| Randomization: |  |  |  |
| Sequence generation | 21a | Who will generate the random allocation sequence and the method used | 9, Supplementary File |
|  | 21b | Type of randomization (simple or restricted) and details of any factors for stratification. To reduce predictability of a random sequence, other details of any planned restriction (e.g., blocking) should be provided in a separate document that is unavailable to those who enroll participants or assign interventions | 9, Supplementary File |
| Allocation concealment  mechanism | 22 | Mechanism used to implement the random allocation sequence (e.g., central computer/telephone; sequentially numbered, opaque, sealed containers), describing any steps to conceal the sequence until interventions are assigned | 9, Supplementary File |
| Implementation | 23 | Whether the personnel who will enroll and those who will assign participants to the interventions will have access to the random allocation sequence | 9, Supplementary File |
| Blinding | 24a | Who will be blinded after assignment to interventions (e.g., participants, care providers, outcome assessors, data analysts) | 9, Supplementary File |
|  | 24b | If blinded, how blinding will be achieved and description of the similarity of interventions | 9, Supplementary File |
|  | 24c | If blinded, circumstances under which unblinding is permissible, and procedure for revealing a participant’s allocated intervention during the trial | 9, Supplementary File |
| **Methods: Data collection, management, and analysis** | | |  |
| Data collection methods | 25a | Plans for assessment and collection of trial data, including any related processes to promote data quality (e.g., duplicate measurements, training of assessors) and a description of trial instruments (e.g., questionnaires, laboratory tests) along with their reliability and validity, if known. Reference to where data collection forms can be accessed, if not in the protocol | 10 |
|  | 25b | Plans to promote participant retention and complete follow-up, including list of any outcome data to be collected for participants who discontinue or deviate from intervention protocols | 10 |
| Data management | 26 | Plans for data entry, coding, security, and storage, including any related processes to promote data quality (e.g., double data entry; range checks for data values). Reference to where details of data management procedures can be accessed, if not in the protocol | Supplementary File |
| Statistical methods | 27a | Statistical methods used to compare groups for primary and secondary outcomes, including harms | 11, Supplementary File |
|  | 27b | Definition of who will be included in each analysis (e.g., all randomized participants), and in which group | 11, Supplementary File |
|  | 27c | How missing data will be handled in the analysis | 11, Supplementary File |
|  | 27d | Methods for any additional analyses (e.g., subgroup and sensitivity analyses) | 11, Supplementary File |
| **Methods: Monitoring** | | |  |
| Data monitoring committee | 28a | Composition of data monitoring committee (DMC); summary of its role and reporting structure; statement of whether it is independent from the sponsor and funder; conflicts of interest and reference to where further details about its charter can be found, if not in the protocol. Alternatively, an explanation of why a DMC is not needed | Supplementary File |
|  | 28b | Explanation of any interim analyses and stopping guidelines, including who will have access to these interim results and make the final decision to terminate the trial | Supplementary File |
| Trial monitoring | 29 | Frequency and procedures for monitoring trial conduct. If there is no monitoring, give explanation | Supplementary File |
| **Ethics** | | |  |
| Research ethics approval | 30 | Plans for seeking research ethics committee/institutional review board approval | 15 |
| Protocol amendments | 31 | Plans for communicating important protocol modifications to relevant parties | 14 |
| Consent or assent | 32a | Who will obtain informed consent or assent from potential trial participants or authorized proxies, and how | 15 |
|  | 32b | Additional consent provisions for collection and use of participant data and biological specimens in ancillary studies, if applicable | 15 |
| Confidentiality | 33 | How personal information about potential and enrolled participants will be collected, shared, and maintained in order to protect confidentiality before, during, and after the trial | 15 |
| Ancillary and post-trial care | 34 | Provisions, if any, for ancillary and post-trial care, and for compensation to those who suffer harm from trial participation | Supplementary File (Intervention Manual) |

*We strongly recommend reading this checklist in conjunction with the SPIRIT 2025 Explanation and Elaboration and the SPIRIT 2025 Expanded Checklist for important clarifications on all the items. We also recommend reading relevant SPIRIT extensions. See [www.consort-spirit.org](http://www.consort-spirit.org)

Citation: Chan A-W, Boutron I, Hopewell S, Moher D, Schulz KF, et al. SPIRIT 2025 statement: updated guideline for protocols of randomised trials. BMJ 2025;389:e081477. <https://dx.doi.org/10.1136/bmj-2024-081477>

© 2025 Chan A-W et al. This is an Open Access article distributed under the terms of the Creative Commons Attribution License (<https://creativecommons.org/licenses/by/4.0/>), which permits unrestricted use, distribution, and reproduction in any medium, provided the original work is properly cited.

#### Table S4 Site Collaborative Group Members

| **Title** | **First name** | **Middle name** | **Surname** | **Email address** | **Primary affiliation** | **Secondary affiliation** | **ORCID iD** |
| --- | --- | --- | --- | --- | --- | --- | --- |
| Dr. | Kevin | B | Moore | | Tallaght University Hospital, Dublin, Ireland |  |  |
| Dr | Ronan |  | Canavan | | St Vincent's University Hospital, Dublin, Ireland |  |  |
| Dr | Nigel |  | Glynn | | Mater Misericordiae University Hospital, Dublin, Ireland |  | 0000-0002-9248-8191 |
| Dr | Wan Aizad |  | Wan Mahmood | | St. Columcille's Hospital, Loughlinstown, Dublin 18 |  | 0000-0002-2437-3018 |
| Dr | Siobhán |  | Bacon | | Sligo University Hospital | School of Medicine, University Of Galway |  |
| Dr | Tríona |  | O'Shea | | University Hospital Waterford |  |  |
| Dr | Sinéad |  | Glackin | | Sligo University Hospital | School of Medicine, University Of Galway | 0000-0003-4521-0223 |
| Prof | Shu |  | Hoashi | | Regional Hospital Mullingar | UCD School of Medicine, University College Dublin |  |
| Prof | Diarmuid |  | Smith | | Beaumont Hospital, Dublin | RCSI School of Medicine |  |

1. Schmitt A, Gahr A, Hermanns N, Kulzer B, Huber J, Haak T. The Diabetes Self-Management Questionnaire (DSMQ): development and evaluation of an instrument to assess diabetes self-care activities associated with glycaemic control. *Health Qual Life Outcomes*. 2013;11(1):138. doi:10.1186/1477-7525-11-138
2. Stanulewicz N, Mansell P, Cooke D, Hopkins D, Speight J, Blake H. PAID-11: A brief measure of diabetes distress validated in adults with type 1 diabetes. *Diabetes Res Clin Pract*. 2019;149:27-38. doi:10.1016/j.diabres.2019.01.026
3. EuroQol - a new facility for the measurement of health-related quality of life. *Health Policy*. 1990;16(3):199-208. doi:10.1016/0168-8510(90)90421-9
4. Anderson RM, Fitzgerald JT, Gruppen LD, Funnell MM, Oh MS. The Diabetes Empowerment Scale-Short Form (DES-SF). *Diabetes Care*. 2003;26(5):1641-1642. doi:10.2337/diacare.26.5.1641-a
5. Eitel KB, Roberts AJ, D’Agostino R, et al. Diabetes Stigma and Clinical Outcomes in Adolescents and Young Adults: The SEARCH for Diabetes in Youth Study. *Diabetes Care*. 2023;46(4):811-818. doi:10.2337/dc22-1749
6. Hobbins A, Barry L, Kelleher D, et al. Utility Values for Health States in Ireland: A Value Set for the EQ-5D-5L. *PharmacoEconomics*. 2018;36(11):1345-1353. doi:10.1007/s40273-018-0690-x
7. ***t_0_*** Baseline (Month 1)

   ***t_1_*** Allocation (Month 1)

   ***t_2_*** Clinic Appointment 1 (Month 4-5)

   ***t_3_*** Clinic Appointment 2 (Month 7-8)

   ***t_4_*** Clinic Appointment 3 (Month 11-12)

   ***t_5_*** Follow up (Month 12-15)
8. In this protocol, the Chief Investigator of the D1 Now Definitive RCT will be referred to as the "PI" or "D1 Now PI" to distinguish from the local PIs at each participating centre.
